# Economic Benefits of COVID-19 Screening Tests with a Vaccine Rollout

**DOI:** 10.1101/2021.03.03.21252815

**Authors:** Andrew Atkeson, Michael Droste, Michael J. Mina, James H. Stock

**Affiliations:** Department of Economics, UCLA; Department of Economics, Harvard University; Chan School of Public Heath, Harvard University; Department of Economics and Harvard Kennedy School, Harvard University

**Keywords:** Epidemiological models, macroeconomics, antigen testing

## Abstract

This note estimates the costs and benefits of a nationwide COVID-19 screening testing program in the presence of vaccine distribution. Even for an optimistic vaccine rollout scenario, a well-designed federally-funded screening testing program, coupled with self-isolation of those who test positive, pays for itself in terms of increased GDP and is projected to save 20,000 or more lives. The sooner the testing program is put in place, the greater are its net economic benefits. This note updates the December 9, 2020 version to include updated deaths data, later dates for rolling out the screening testing program, and the spread of more contagious variants such as the B.1.1. 7 variant.

---

Since early in the COVID-19 pandemic, there have been calls for widespread, inexpensive and convenient COVID-19 screening tests as a way to identify infected individuals and control the spread of the virus.^1^ This note reports the results of a macroeconomic cost-benefit analysis of a federally-funded national COVID-19 screening testing program in the presence of a vaccine distributed during 2021.^2^

We consider a vaccine distribution path that matches distribution to date and projects 75% of the population to be vaccinated by August 1, 2021. We consider two testing regimes (10-day and 5-day cadences) and three screening testing regime start dates (Feb. 15, March 1, and March 15, 2021). The rapid test we consider is modeled on Innova Medical Group’s rapid antigen test, for which we assume a specificity of 99.5% and a sensitivity of 90%. We assume that 50% of those testing positive on the screening test receive a confirmatory PCR test, and that the self-isolation adherence rate for those testing positive on the antigen test, but not taking a confirmatory PCR test, is 50%.

Table 1 presents estimates of the economic and mortality impacts of hypothetical screening test programs (relative to a baseline with no additional screening tests). Panel A considers screening test programs with a 10-day cadence; that is, approximately 33 million tests per day. Panel B considers a program with a 5-day cadence, and panel C considers a 3-day cadence. Each row corresponds to a different roll-out date. The screening and diagnostic tests are assumed to cost $5 and $50, respectively. All results are shown relative to a benchmark of no federal screening testing program.

**Table 1.** Economic and mortality impacts of screening programs.

| Roll-out | Additional testing costs (\$B) | Additional GDP (\$B) | Additional federal receipts (\$B) | Deaths averted (Thou) |
| --- | --- | --- | --- | --- |
| *A. 10-day screening testing* |  |  |  |  |
| Feb 15 | 3.29 | 27.21 | 2.96 | 48.70 |
| Mar 1 | 2.88 | 16.86 | 1.83 | 38.58 |
| Mar 15 | 2.47 | 8.33 | 0.91 | 28.32 |
| *B. 5-day screening testing* |  |  |  |  |
| Feb 15 | 5.93 | 38.46 | 4.18 | 73.21 |
| Mar 1 | 5.17 | 25.19 | 2.74 | 58.57 |
| Mar 15 | 4.83 | 13.02 | 1.42 | 42.40 |
| *C. 3-day screening testing* |  |  |  |  |
| Feb 15 | 8.79 | 46.19 | 5.02 | 91.01 |
| Mar 1 | 7.68 | 31.80 | 3.46 | 73.93 |
| Mar 15 | 6.52 | 17.15 | 1.87 | 55.02 |

*Note: Figures are relative to a baseline scenario a uniform population daily screening rate of 1/328, about one million tests per day. Simulation period runs through Jan 1, 2023. Screening test sensitivity is 90% and specificity is 99.5%. Adherence to self-isolation among those testing positive is 50% for screening test alone.*

The hypothetical screening test programs described in Table 1 avert between 28,000 and 91,000 deaths in our model. The reduction in the prevalence of the virus leads to an increase in GDP between 8 and 46 billion dollars. The impact of the testing program depends heavily on the roll-out date: programs that start earlier avert more deaths and lead to a larger increase in GDP (relative to a no-screening counterfactual).

Broadly, our results suggest the following three conclusions:

1. Across all scenarios, the increase in GDP resulting from the testing program ranges from 2 to 8 times the incremental cost of the tests.
2. The timing of the introduction of the testing program has a large impact on the program’s net benefits. For example, for the cases in Table 1, starting the testing program on February 15 instead of March 1 saves an additional 10,000 lives for a program with a 10-day cadence, and 15,000 lives for a program with a 5-day cadence.
3. In all scenarios, a significant share of the program costs are made up for by additional federal receipts induced by increased business activity when viral prevalence is low.

These calculations assume homogenous distribution of tests to all those who have not been vaccinated. A targeted testing regime could have different effects, for example if testing were focused on individuals with the most contacts, virus prevalence and deaths might be less than those projected in Table 1. Because of the endogenous response of economic activity to deaths, the resulting lower death rate would lead to higher levels of GDP than those reported here.

The assumptions used in this calculation are conservative in several ways. First, the model assumes random testing and homogenous contacts within age groups. In reality, different groups have different contact rates, and if the testing is targeted to those with the highest contacts, the effects of the same number of tests, but targeted, would be greater than reported here. Second, we assume that the vaccine efficacy rate is also the rate of reduction of transmissibility. If the vaccine allows mild cases with transmission, then it will take longer to control the pandemic and the testing program will be more valuable. Third, we allow for the B.1.17 variant to be more contagious but not more deadly, nor do we allow for more deadly variants, nor do we allow for reduced vaccine efficacy against those variants. If any of those assumptions are wrong, the pandemic would take longer to eliminate and testing would be more valuable.

## Model and assumptions

The ADMS model is a 66-sector national economic model linked through behavioral feedback to a 5-age national SIR model that includes susceptible, exposed, infected, recently recovered, fully recovered, and deceased compartments. All but the deceased compartments are differentiated by testing status (awaiting a screening test result, awaiting a diagnostic test result, or positive and self-isolating). Screening tests are given randomly to the non-symptomatic. Diagnostic tests are given to the symptomatic and are offered as a confirmatory test to those who test positive on the screening test. Because the screening test has imperfect specificity, the positive predictive value of a screening test can be low (typical values are 5%-25% for the parameter values considered here), so adherence to self-isolation is modeled as being partial in response to a positive screening test. The screening (antigen) test is assumed to be sensitive only to those infected, whereas the diagnostic (PCR) test is taken to be sensitive both to the infected and to the recently recovered. For additional discussion, see ADMS (2020a).

We extend the ADMS model to incorporate an exogenous path for vaccination. Vaccination is modeled as occurring randomly and independently of the infection status (susceptible, exposed, etc.), prior test results, and age. Vaccination is modeled as binary and completed so that a vaccinated individual is immune with probability calibrated to estimates of efficacy derived from the Moderna and Pfizer Phase 3 clinical trials. It is not known whether a given individual becomes immune, and if she does not, she remains susceptible and active if she was susceptible prior to vaccination. Those vaccinated are removed from the screening and diagnostic testing programs.

Technical updates to the model include the addition of seasonality, re-estimation through Nov. 10, 2020, and updated estimates of the feedback rule. These updates are summarized in the Appendix.

### Testing, Vaccination, and Masking Scenarios

#### Testing

The baseline testing scenario is the current level of screening testing (approximately 1 million tests per day) and the current level of diagnostic testing. The federal screening testing programs make screening testing available to all non-vaccinated individuals at a specified rate (once every 5 or 10 days). Those who test positive on the screening test are offered a (free) confirmatory PCR test; we assume that 90% of screening-positives take a confirmatory PCR test. All additional testing costs are paid by the federal government. The screening test is assumed to exhibit 99.5% specificity and 90% sensitivity, and a marginal cost of $5 per test. Note that in our model, the concept of test sensitivity corresponds to the share of those who are truly infectious who would test positive on a screening test. In practice, true infectiousness at a point in time is difficult to measure, and there is a large body of evidence suggesting that the gold-standard PCR tests can detect viral genetic sequences even after an individual has ceased to become infectious. We assume 50% adherence for those testing positive on the screening test and not taking a confirmatory PCR test. We assume that PCR confirmatory tests exhibit 99.9% sensitivity, 99.7% specificity, and a marginal cost of $50/test. The federal screening testing program is shut down when daily deaths fall below 100.

#### Vaccination

Our model takes as given a forecasted path for the distribution of vaccines to the U.S. population. At the time of writing, this forecast is subject to uncertainty for several reasons: (1) there is uncertainty in the number of distinct vaccines that will be available in the United States; (2) the timetable for the delivery of vaccines purchased by the United States remains uncertain; and (3) voluntary uptake in the population may limit vaccination when supply catches up to demand. At the time of writing, two vaccines (produced by Pfizer and Moderna) have received emergency use authorization for use in the United States. In addition, a one-dose vaccine produced by Johnson and Johnson is expected to receive an emergency use authorization in a matter of weeks. Collectively, these companies expect to distribute 400 million two-dose vaccines (200 million each of those produced by Pfizer and Moderna) and 100 million one-dose vaccines (from Johnson and Johnson) by the end of June – sufficient to fully vaccinate nearly the entire U.S. population.

Even if these production and distribution targets are met, it remains uncertain how quickly vaccines distributed will make their way into the arms of patients. Current polling data suggests that the take-up could be low: for example, a Gallup poll in November reported only 58% of American adults would be willing to take the vaccine. Acceptance could increase as the vaccine is observed to be safe and effective, or it could fall if safety concerns arise. Vaccination could also be slowed if supply chain or administrative disruptions occur.

Figure 1 documents the cumulative share of the population who is vaccinated at each point in time under our vaccine distribution timetable. For simplicity, we do not distinguish between available vaccines. We also assume the vaccine exhibits 90% efficacy in preventing infection.

**Figure 1.**
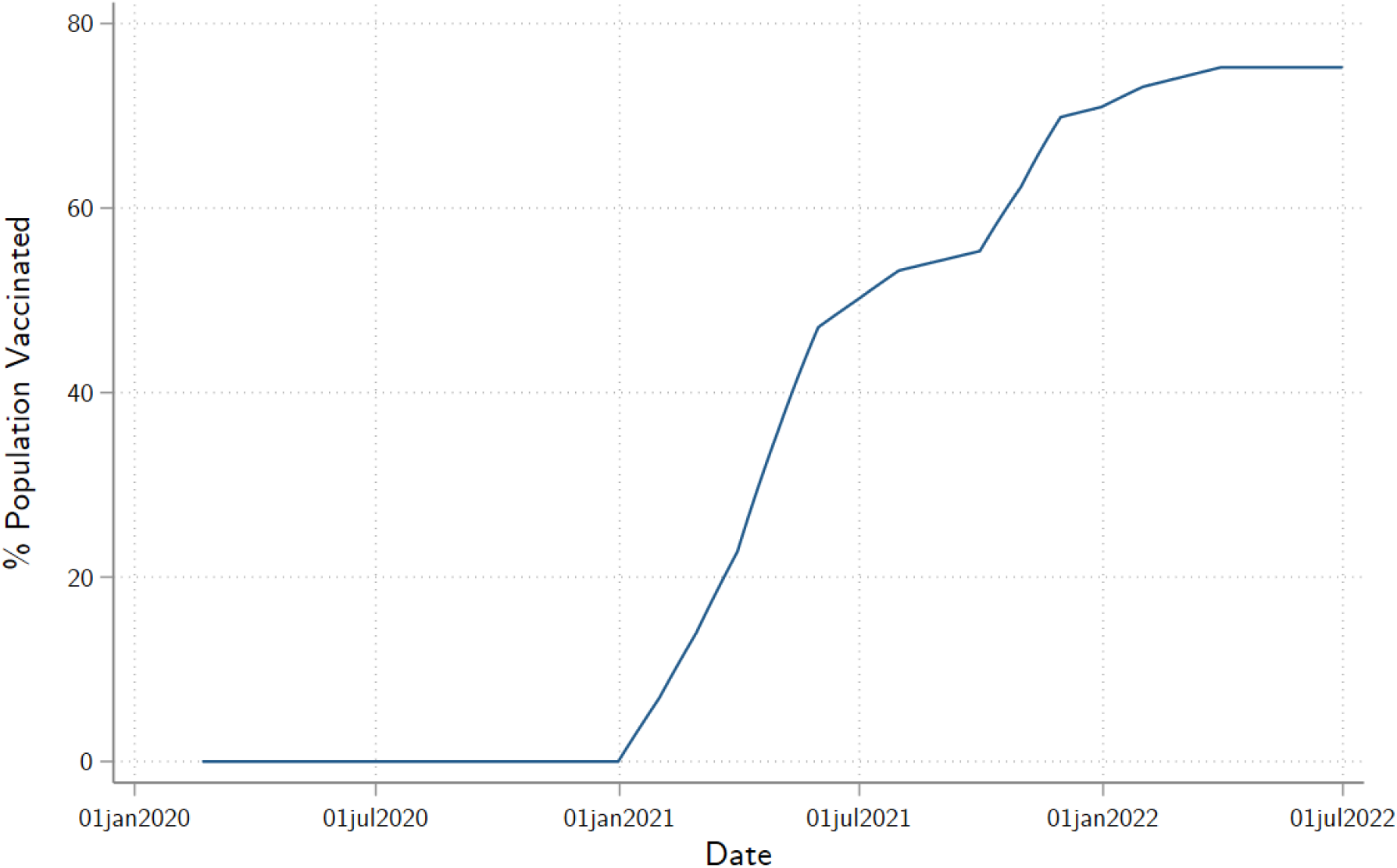
Vaccination Forecast Path.

### Cost-Benefit Estimates

We compute two measures of the economic benefits of screening test programs: incremental GDP and incremental federal government revenues. Monetary values are in 2020 dollars. Revenues flowing from an increase in GDP are computed following the Congressional Budget Office (Russek and Kowalewski (2015, Table 3) and CBO (2019)).

## Data Availability

All data used in this paper is publicly available.

## Appendix: Technical Updates and Parameter Values

This appendix summarizes three technical updates of the model, relative to ADMS.

First, the model was updated to include seasonality in the transmissibility of the virus. The literature on COVID seasonality is limited because COVID has not existed for a full seasonal cycle. IHME (2020) calibrated COVID seasonality using pneumonia seasonality. We rely instead on regression estimates of COVID seasonality in Tzampoglou and Loukidis (2020), who use country-level data from March through May 2020. Their estimates imply fluctuations in deaths of 21% to 32% as a result of temperature variation across the full range of cross-country temperatures. We adopt the approximate midpoint of this range and use a 25% full seasonal fluctuation in transmissibility, which we approximate as a seasonal cosine peaking on January 1, a pattern consistent with Merow and Urban (2020).

Second, we reestimated the model using daily U.S. deaths data (7-day moving average) from March 15, 2020 through Nov. 10, 2020. The simulation period runs from Nov. 7, 2020 through July 1, 2022. The ADMS model uses a nonparametric flexible functional form to fit unobserved self-protective behavior such as masking and social distancing, specifically on a Type-II cosine expansion with three terms. The ADMS parameter estimation sample was March 15 – June 12, 2020. For the longer sample used here, we added two more terms to the Type-II cosine expansion. Also, the ADMS model calibrated protections for the elderly, however the longer sample afforded sufficient data to estimate a separate factor multiplying the cosine expansion to apply to the oldest age group, 75+. In all the model used here has 8 estimated parameters: the number of infections on February 21, 2020, the initial transmissibility rate per adult contact (equivalently, the initial *R*_0_), the five parameters of the cosine expansion, and the elderly protection factor.

Third, the behavioral feedback rule was updated. ADMS considered two behavioral response rules from deaths to economic activity: the calibrated rule used in Baqaee, Farhi, Mina, and Stock (2020) and a simpler rule estimated by Arnon, Ricco, and Smetters (2020). Both those feedback rules are based on data from Spring 2020. A plausible concern is that the response of economic activity to the prevalence of the virus diminished as individuals adapt and experience lockdown fatigue. Here, we therefore adopt a behavioral rule in which economic activity depends on the current death rate (as in Arnon, Ricco, and Smetters (2020)), where the feedback parameter is estimated so that an exogenous disappearance of the virus would bring labor hours, as a fraction of potential, to a specified level. ^3^

## Acknowledgements

Droste and Stock acknowledge research support under NSF RAPID Grant SES-2032493.

## Footnotes

1 See for example Gottlieb et al. (2020), National Governors’ Association (2020), The Conference Board (2020), Romer (2020), Rockefeller Institute (2020), Silcox et al (2020), and Kotlikoff and Mina (2020).

2 This analysis updates Atkeson, Droste, Mina, and Stock (2020b) (ADMSb), which in turn was based on Atkeson, Droste, Mina, and Stock (2020a) (ADMSa). ADMSa undertook a macroeconomic cost-benefit analysis of a national testing program with partial adherence, counterfactually introduced in the summer of 202 0 in the absence of a vaccine. ADMSa,b used a 66-sector, five-age US national linked epidemiological-economic model that incorporates an economic behavioral response to the COVD-19 death rate. Here, we update ADMSb to incorporate new data on the winter wave, to consider a testing program rolled out in February or March 2021, and to include the spread of more contagious variants (with timing based on CDC projections), in conjunction with vaccine distribution during 2021.

3 Specifically we assume a post-pandemic level of labor hours at 96.4% of potential, which we compute from the Hall-Kudlyak (2020) estimate of the unemployed without jobs plus the change in the labor force participation rate from February through November 2020.

